# Heritability and polygenic load for combined anxiety and depression

**DOI:** 10.1101/2024.01.31.24302045

**Authors:** Fara Tabrizi, Jörgen Rosén, Hampus Grönvall, Victor Rahimzadeh William-Olsson, Erik Arner, Patrik KE Magnusson, Camilla Palm, Henrik Larsson, Alexander Viktorin, Jens Bernhardsson, Johanna Björkdahl, Billy Jansson, Örjan Sundin, Xuan Zhou, Doug Speed, Fredrik Åhs

**Author notes:** Corresponding author, Department of Psychology & Social Work, Mid Sweden University Kunskapens vag 8, Ostersund Room: P1517.

## Abstract

Anxiety and depression commonly occur together resulting in worse health outcomes than when they occur in isolation. We aimed to determine whether the genetic liability for combined anxiety and depression was greater than when anxiety or depression occurred alone. Data from 12,558 genotyped twins (ages 38-85) were analysed, including 1,986 complete monozygotic and 1,809 complete dizygotic pairs. Outcomes were prescription of antidepressant and anxiolytic drugs, as demined by the World Health Organization Anatomical Therapeutic Chemical Classimication System (ATC) convention, for combined anxiety and depression (*n* = 1054), anxiety only (*n* = 744), and depression only (*n* = 511). Heritability of each outcome was estimated using twin modelling, and the inmluence of common genetic variation was assessed from polygenic scores (PGS) for depressive symptoms, anxiety, and 40 other traits. Heritability of combined anxiety and depression was 79% compared with 41% for anxiety and 50% for depression alone. The PGS for depressive symptoms likewise predicted more variation in combined anxiety and depression (adjusted odds ratio per *SD* PGS = 1.53, 95% CI = 1.43-1.63; Δ*R*^2^ = .031, ΔAUC = .044) than the other outcomes, with nearly identical results when combined anxiety and depression was demined by International Classimication of Diseases (ICD) diagnoses (adjusted odds ratio per *SD* PGS = 1.70, 95% CI = 1.53-1.90; Δ*R*^2^ = .036, ΔAUC = .051). Individuals in the highest decile of PGS for depressive symptoms had over 5 times higher odds of being prescribed medication for combined anxiety and depression compared to those in the lowest decile. We conclude that genetic factors explain substantially more variation in combined anxiety and depression than anxiety or depression alone.

---

Anxiety– and mood disorders are the most commonly occurring psychiatric disorders in the general population, with lifetime prevalence estimates of up to 33.7% and 14.7%, respectively (1,2). Symptoms of these disorder categories most often occur together, as evident by nearly half of adults with anxiety also reporting depressive symptoms(3) and two-thirds of patients with a primary diagnosis of major depressive disorder (MDD) also displaying pathological levels of anxiety (4–6). It is of importance to distinguish between combined anxiety-depression and anxiety or depression occurring alone because the combination has worse health outcomes, entails greater risk of suicide, and is more resistant to treatment (7–9). Despite being highly debilitating and prevalent, the heritability of this phenotype combination is largely unknown. Hence, in the current study we aimed to determine whether the genetic contribution to combined anxiety and depression differs from depression or anxiety alone.

Meta-analyses report twin-heritability in the range of 30-50% for separate occurrences of anxiety– and depressive disorders (10,11), indicating a moderate heritable component. Although the shared heritability between these disorder categories is known to be large (12,13), twin-studies on combined anxiety and depression are scarce. In a rare instance, Guffanti et al. (14) estimated the heritability for sequential comorbidity of anxiety disorders and MDD in the range of 53-57% using a relatively small sample of 545 high-risk participants from 65 multigenerational families. The findings by Guffanti et al. suggest that the combination of anxiety and MDD might be more heritable than when each disorder occur alone, but the small sample size together with the probable oversampling of cases warrants replication before firm conclusions can be drawn.

Anxiety– and depressive disorders have also been predicted from polygenic scores (PGS) that are based on genomic variations associated with phenotypes in independent discovery samples (15). To date, anxiety PGSs explain about 0.5% of variance when predicting anxiety disorders in independent cohorts (16,17), while depression PGSs explain 0.27-2.2% of variance in depression (18,19). Tentative evidence further suggests that individuals with comorbid anxiety have higher PGSs for a broad depression phenotype (odds ratio per *SD* PGS = 1.17) compared to participants meeting only MDD criteria (20). These results have been corroborated by recent findings, indicating that depression with comorbid anxiety, as defined by ICD codes, is associated with higher polygenic load for several psychopathological traits in comparison with depression without anxiety (21). Although these reports suggest increased genetic liability for combined anxiety and depression compared to when anxiety or depression occur alone, the limited number of studies and the differences in how cases have been ascertained, make generalizable conclusions premature.

Electronic health-record based studies of genetic influences on anxiety and depressive disorders commonly depend on diagnoses from specialist care. This could potentially induce a bias in estimates of genetic liability as most individuals seeking treatment for anxiety or depression initially consult a general practitioner in primary care, and often receive a drug prescription from these practitioners without referral to a specialist. To include patients seeking care for anxiety (*Anxiety-only*), depression (*Depression-only*), or both (*Combined* anxiety and depression) from both primary and specialist care, we here used an algorithm to search the Swedish Prescribed Drug Register for information about the symptoms for which drugs were prescribed. This so-called dosage text is written by the GP and contains the main indication for the drug prescription. To validate the ascertainment of cases, we compared which drug classes were most commonly prescribed for each symptom category (Anxiety-only, Depression-only, Combined anxiety and depression). In addition to symptom categories based on drug prescription, outcome groups were also defined by International Classification of Diseases (ICD) diagnoses that only included patients referred to specialist care. This allowed us to evaluate whether genetic influences on outcome categories were dependent on the definition of cases (ICD diagnoses vs. drug prescription information). Notably, the term combined here refers to having been prescribed drugs for symptoms of anxiety and depression or diagnosed with a depression and an anxiety diagnosis within a given time period without necessarily being simultaneously present at any point in that time period (22).

For the purpose of this study, we constructed PGSs for 6 different traits focusing on anxiety, depression, schizophrenia, and neuroticism, while also using 36 predefined PGSs from the polygenic index repository (23). We then sought to understand how various PGSs could predict categories of individuals prescribed medication for symptoms of anxiety, depression, or both.

This analysis could reveal whether a specific PGS shows an advantage in predicting symptom– or disorder categories compared to other PGSs. It would also inform on whether one indication for drug prescription (i.e., one symptom category) has more genetic liability over others. The analyses were then repeated with outcomes defined by ICD diagnoses. By using a sample of monozygotic and dizygotic twins from the Swedish Twin Registry, we could also determine if there was a correspondence between genetic liability of outcomes estimated from PGSs and heritability using classic twin modelling.

Another aim was to determine the effect of having extremely high or low PGSs on anxiety, depression or combined anxiety and depression. We estimated risk carried by extreme PGSs by comparing individuals in each decile with those in the lowest decile of the PGS distribution to determine the utility of PGSs regarding risk stratification.

## Method

### Study population

STAGE and YATSS are population cohorts of Swedish twins born between 1959-1985 and 1986-1992, respectively, that are part of the Swedish Twin Registry (24,25). The STAGE study invited 25,364 twins to donate saliva for genotyping in 2005-2006, and 6,218 twins from YATSS were invited in 2013-2014. Genotyping was performed on the 650K Illumina Global Screening Array BeadChip for both cohorts in 2017-2019, resulting in a final sample of 13,699 twins with genotype data available for 12,792 individuals. STAGE responders were representative of the Swedish population in terms of educational attainment and family background characteristics (26).

## Measures

### Anxiety and Depression Outcomes

We obtained drug prescription data for the entire study population from the Swedish Prescribed Drug Register (27) through the Swedish Twin Registry (STR). We included N06AA non-selective monoamine reuptake inhibitors, N06AB selective serotonin reuptake inhibitors (SSRIs), N06AF monoamine oxidase inhibitors (non-selective), N06AG monoamine oxidase A inhibitors, N06AX other antidepressants (which include certain selective serotonin and noradrenaline reuptake inhibitors as well as other drugs), N05BA benzodiazepine derivatives, N05BE azaspirodecanedione derivatives, R06A antihistamines for systemic use, C07AA beta blocking agents (non-selective), and N03AX other antiepileptics (i.e., pregabalin) based on their Anatomical Therapeutic Chemical code (ATC) (28). For the above-mentioned ATC-categories, we employed an algorithm that searched through the prescription text, also known as the dosage text, written by the prescribing physician. If the text included any of the words “worry”, “anxiety”, “anxiety issues” (one word in Swedish “orosbesvär”), or “panic”, the drugs were classified as prescribed for symptoms of anxiety. For depression, one of the words “depression”, “depressive”, “dejection” (i.e., “nedstämdhet” in Swedish), or “mood-enhancing” had to be included in the text. The text-searching algorithm resulted in two columns for each drug, one column for symptoms of anxiety and one for symptoms of depression, where individuals were coded as 1 or 0 based on if the drug had been prescribed with or without the relevant target words. Those without prescriptions of the target drugs were coded as NA and included as controls. All drug prescriptions were treated as life-time events and there was no distinction made between individuals prescribed drugs for anxiety and depression simultaneously, meaning the terms occurred in conjunction in the dosage text, or individuals prescribed medication for anxiety on one occasion and depression on another, meaning that the terms occurred in two or more separate dosage texts. Based on the results from the text-searching algorithm, we classified individuals into one of four categories, (1) if the text mentioned both anxiety and depression they were included in the symptom category Combined anxiety and depression; (2) if the text mentioned anxiety, but not depression, participants were included in the category Anxiety-only; (3) if the text mentioned depression, but not anxiety, they were included in the category Depression-only; (4) if the text did not mention anxiety or depression they were categorized as *Medication-only*. The last group was included as the target drugs can be prescribed for other indications than anxiety or depression (e.g., otherwise unspecified sleep disorder, premenstrual syndrome, or adjustment disorder) and physicians sometimes omit stating the indication that the medication is prescribed for. These symptom categories were used as outcomes in subsequent statistical analyses, controls comprised those not included in any of these categories.

We also obtained diagnoses from the National Patient Register, which are assigned by an attending physician based on the International Classification of Diseases, eighth revision (ICD-8) (1969–1986), ninth revision (ICD-9) (1987–1996), or tenth revision (ICD-10) (1997-present). Similar to drug prescriptions, diagnoses were defined as life-time events and treated as binary variables. Participants were categorized in the depression diagnosis group if they had any depression-related ICD code, including 3004 (ICD-8), 300E, 311 (ICD-9), and F32-F39 (ICD-10). Anxiety diagnosis was defined as having at least one of the following codes, 300 (excluding 3003, 3004) (ICD8), 300 (excluding 300D, 300E) (ICD-9), and F40-41 (excluding F41.2), F44-45, F48 (ICD-10). The F41.2 code was excluded from the anxiety diagnosis group, since it refers to having symptoms of both anxiety and depression, but where neither is clearly predominant. Participants with an F41.2 code were grouped in a third category (combined depression and anxiety diagnoses), which also included those that had both a depression-related and an anxiety-related ICD code. These 3 groups (Combined anxiety and depression, Anxiety-only, Depression-only) were used as separate outcomes in subsequent analyses, in which controls included participants that neither had a diagnosis nor been prescribed any antidepressants or anxiolytic drugs.

### Polygenic Scores (PGS)

A total of 42 PGSs were evaluated. Thirty-six of these came from the polygenic index (PGI) repository, which contains scores derived from large-scale GWASs conducted in 23andMe, UK Biobank (29), as well as other available datasets. For each phenotype, PGSs have been constructed in 11 repository cohorts including the STR, with a leave-one-out approach to distinguish the discovery sample from the target cohort. Hence, PGSs in STAGE and YATSS were created from GWAS summary statistics that exclude these datasets, which allows for unbiased targeting of phenotypes in our sample to test the predictive performance of PGSs from the PGI-repository. Information on quality control and PGS construction have been reported in detail elsewhere (23).

In addition to the 36 PGSs from the PGI-repository, we constructed 6 PGSs based on Lifetime Anxiety Disorder diagnosis (LAD; cases = 31977; controls = 82114), Major Depressive Disorder (MDD; cases = 170756; controls = 329443), schizophrenia (SCZ; cases = 67390; controls = 94015), Neuroticism scores (NEURO; *n* = 323415), the Generalized Anxiety Disorder 7-item scale (GAD-7; *n* = 126175), and the Patient Health Questionnaire-9 (PHQ-9; *n* = 126733). The LAD-PGS was based on summary statistics from the meta-analysis performed by Purves et al. (17), the MDD-PGS was based on summary statistics published in Wray et al. (19) and Howard et al. (30), and the SCZ-PGS was created from summary statistics obtained from Ripke et al. (31). The NEURO, GAD7, and PHQ9 PGSs were all based on individual level data from the UKB (29,32). Ambiguous SNPs (alleles A/T or C/G), trivial SNPs (showed no variation across UKB) and SNPs with a minor allele frequency (MAF) < 0.01 were all excluded. We used the LDAK software (33) with the BLD-LDAK heritability model (34), where contribution of a SNP depends on MAF, LD and functional annotations. PGS construction was performed with LDAK-Bolt-Predict for individual-level data as well as LDAK-BayesR-SS for summary statistics, and was limited to SNPs that overlapped between cohort genotype, UKB reference panel and GWAS summary statistics. Finally, we converted all PGSs to *z*-scores for interpretation purposes.

## Statistical analyses

### Twin heritability

Twin heritability (*h*^2^_TWIN_) was estimated by fitting structural equation models that decomposed variance in the outcome measures into additive genetic (*A*), common-environmental (*C*) and unique-environmental variance (*E*) as well as with models that only estimated the A and E components. These models are based on the assumptions that Monozygotic (MZ) and Dizygotic (DZ) twin pairs share common environmental effects to the same extent and that MZ twins share 100% of their segregating alleles, while DZ twins share 50% of their segregating alleles. Thus, the additive genetic effects are shared with correlation equal to 1 between MZ pairs and correlation 0.5 between DZ pairs. Analyses were initially performed using only same sex twin pairs and were then repeated including DZ pairs with different sexes in addition to same sex twin pairs. Due to the case/control design (i.e., binary traits as outcome measures), we used the liability threshold model, which assumes a latent bivariate normal distribution of the observed dichotomous variables (35,36). Model fit was assessed by likelihood ratio tests (LRT) and Akaike’s information criteria (AIC). All twin models included sex and age as covariates. Twin heritability was analysed with the mets R-package (37).

### PGS prediction of anxiety and depression outcomes

To determine effect sizes of PGSs in predicting each of the outcomes based on prescription data (Combined anxiety and depression, Anxiety-only, Depression-only, Medication-only) or diagnosis groups (Combined depression and anxiety diagnoses, anxiety diagnosis, depression diagnosis), we used logistic regression. Effect sizes are presented as adjusted odds ratios (OR) per *SD* PGS, with 95% confidence intervals (95% CI) for each outcome. Pseudo-*R*^2^ (Nagelkerke *R*^2^) was estimated for the full model (all covariates including PGS) and the base model (not including PGS). The latter included sex, age, zygosity (to correct for clustering of family members), the first 20 ancestral principal components (to control for population stratification), and cohort (STAGE, YATSS) as covariates. Proportion of predicted variance was calculated as the difference between the two pseudo-*R*^2^ estimates (*ΔR*^2^). We also calculated area under the Receiver operating characteristics curve (AUC) between the base model and the full model including the best performing PGS.

We next included all PGSs in one regression model to assess the ability of each score in predicting the outcomes while controlling for the other PGSs, and evaluated the *ΔR*^2^ to assess how much variance in outcomes could be predicted when all genetic indices were included in the same model. Collinearity diagnostics showed that all variables had a variance inflation factor and tolerance statistic well below 10 and above 0.1, respectively, indicating no significant multicollinearity.

To describe associations across the PGS distribution with each outcome, we first selected the PGSs that had the overall strongest association with the outcomes according to their OR and *p*-value, and divided the sample in deciles based on their PGS. Using logistic regression, we computed the association between each outcome and the highest PGS decile compared with the remaining 80% of the sample (leaving out the bottom 10%). The same analysis was performed for the lowest decile (leaving out the top 10%). These analyses inform on how groups with extreme scores compare with those that have scores closer to the mean. To assess the incremental increase in association between PGS and each outcome across deciles, separate regression analyses were performed for each decile using the lowest as reference.

All *p*-values were corrected for multiple comparisons using the Benjamini-Hochberg procedure (38) for False Discovery Rate (FDR < .05).

## Results

### Sample characteristics

Data on drug prescription were retrieved for 13699 individuals, where genotype information was available for 12792 participants (see Table 1 for sample characteristics). Due to missing data for sex and age, the final sample for PGS-predictions using regression models with covariates included 12558 individuals, whereas twin heritability analyses were based on 1986 complete MZ-pairs and 1809 DZ-pairs. The control group for analyses of prescription-based outcomes include all individuals without any prescriptions of the target drugs, while analyses of diagnostically defined outcomes included those without drugs or diagnoses in the control group (see supplemental material for the exact *n* in each analysis).

**Table 1.**
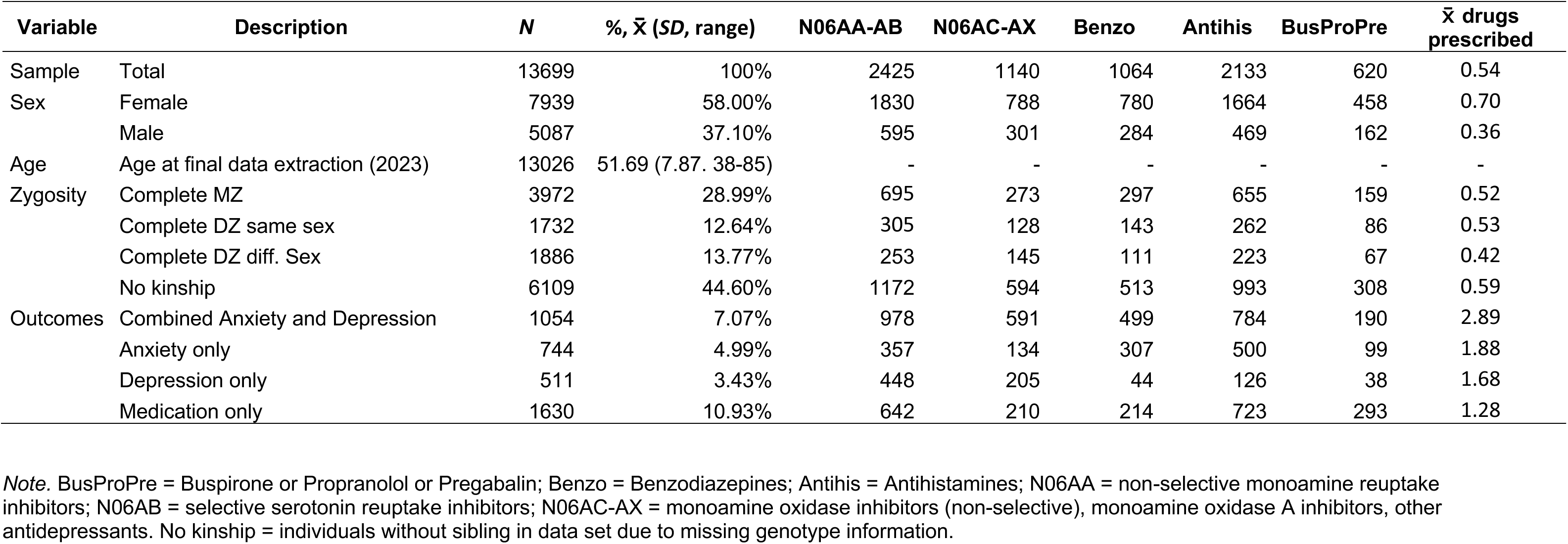
Sample Characteristics.

To compare patterns of prescriptions across outcome groups, we computed the proportion of individuals in each group that were prescribed medications from the drug classes N06AA-AB, N06AC-AX, benzodiazepines, antihistamines and Buspirone/Propranolol/Pregabalin (BusProPre) (Fig. 1). In the Anxiety-only category, drugs from the class antihistamines were most frequently prescribed, followed by N06AA-AB and benzodiazepines. In contrast, the Depression-only group had the lowest proportion of prescriptions from the benzodiazepine class compared to any of the other groups. Instead, N06AA-AB was the most commonly prescribed drug class for this group followed by N06AC-AX. The Combined anxiety and depression group had high prescription rates of drugs from the classes N06AA-AB and antihistamines as well as benzodiazepines and N06AC-AX. Notably, individuals in the Combined symptom category were on average prescribed drugs from more drug classes (2.89) compared to any of the other outcome groups (1.28-1.88). The Medication-only group had the lowest rates of prescriptions (1.28) with antihistamines being the most frequently prescribed, followed by N06AA-AB and BusProPre. These results indicate that the prescription text that was used for categorizing individuals into the outcome groups was associated with different treatment decisions.

**Figure 1.**
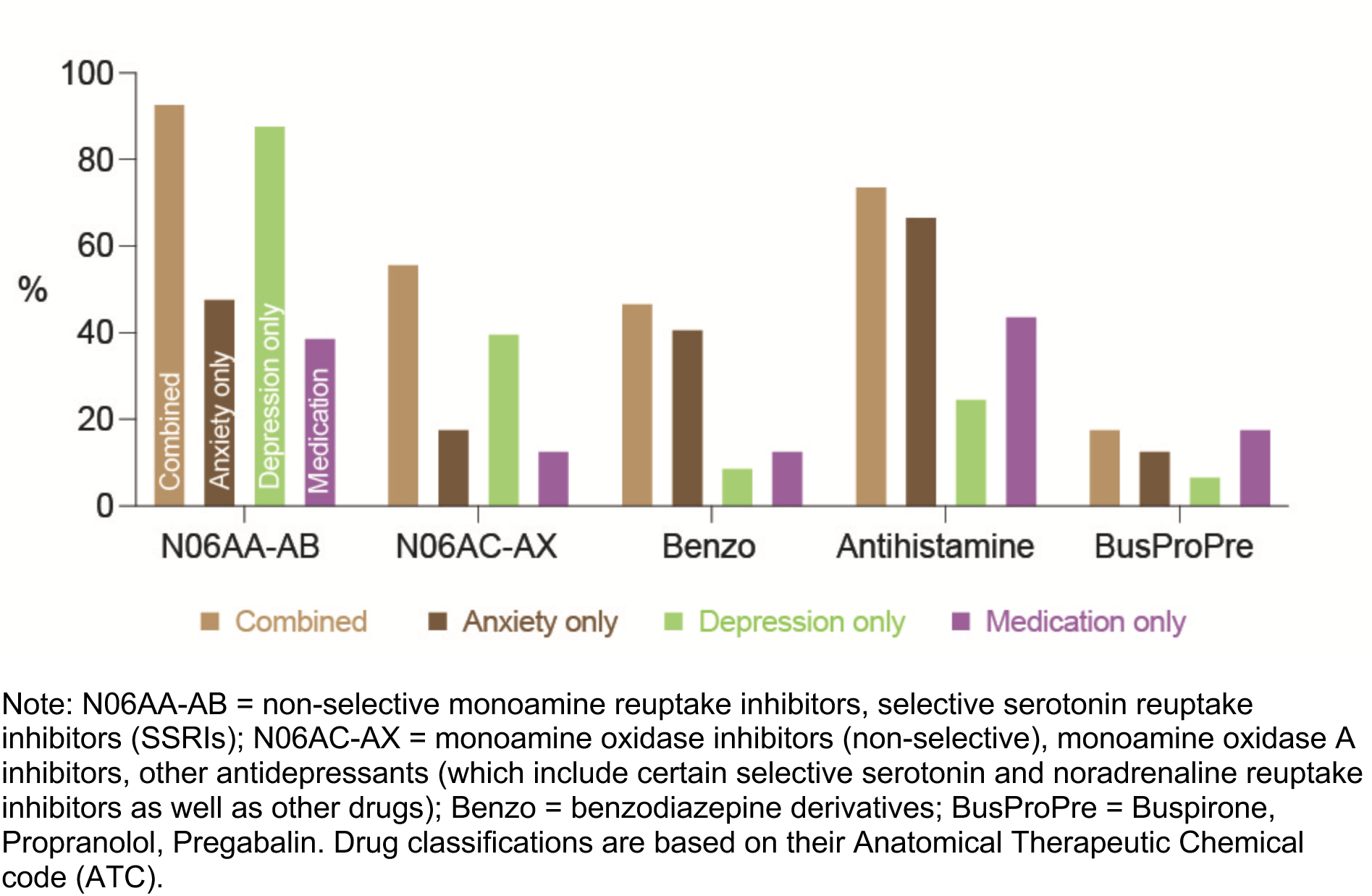
Proportion of outcome groups with prescription divided on by drug class.

To assess diagnoses of anxiety and depressive disorders, we retrieved data from the National Patient Registry. Out of the 1029 participants with an ICD code associated with these disorders, 379 had only a depression diagnosis, 283 had only an anxiety diagnosis, and 367 had combined depression and anxiety diagnoses. The group with combined anxiety and depression diagnoses consisted of 173 individuals that had both a depression– and an anxiety-related ICD code, 40 individuals with an F41.2 (mixed anxiety and depressive disorder) diagnosis, 50 individuals with F41.2 and an anxiety ICD code, 27 individuals with F41.2 and a depression ICD code, and 77 individuals with F41.2 plus a depression and an anxiety ICD code. As expected, the number of individuals with diagnoses were fewer than the number of individuals with prescriptions, as most individuals receiving prescriptions from primary care are not referred to a specialist that enters a diagnosis in the Swedish patient registry.

### Twin-based heritability of anxiety and depression

The additive genetic influence on having been prescribed medication for combined anxiety and depression was *h*^2^_TWIN_ = 0.79 (95% CI = 0.71-0.86). Lower additive genetic effects were found for having been prescribed medication for Anxiety-only (*h*^2^_TWIN_ = 0.41, 95% CI = 0.23-0.60) or Depression-only (*h*^2^_TWIN_ = 0.50, 95% CI = 0.26-0.75). Further, having been prescribed the same drugs, but without an indication of anxiety or depressive symptoms, resulted in a heritability estimate of *h*^2^_TWIN_ = 0.29 (95% CI = 0.17-0.40). See supplementary Table S1 for twin-model estimates.

## Prediction of anxiety and depression from PGSs

### Regression models with single PGS predictors

Figure 2 shows which PGSs that were significantly associated with the different outcomes, and the strength of associations in terms of OR per *SD* PGS. When modelled as individual predictors, 27 out of the 42 PGSs evaluated (36 from the PGI repository and 6 scores computed for the purpose of this study) showed a significant increase in predicting drugs prescribed for Combined anxiety and depression compared with the base model (base model *R*^2^ = .041, AUC = .632). Out of these 27 PGSs, the PGS for depressive symptoms from the PGI repository (DEP-PGS) was the strongest predictor (Δ*R*^2^ = .031, ΔAUC = .044, OR = 1.53, 95% CI = 1.43-1.63) followed by the PGS for major depressive disorder (MDD-PGS) computed from Wray et al. (19) using LDAK (Δ*R*^2^ = .020, ΔAUC = .032, OR = 1.39, 95% CI = 1.30-1.49), PGS for Subjective Wellbeing (SWB-PGS; Δ*R*^2^ = .018, ΔAUC = .028, OR = 0.73, 95% CI = 0.68-0.78), Lifetime Anxiety Disorder (LAD-PGS) computed with LDAK (Δ*R*^2^ = .015, ΔAUC = .022, OR = 1.33, 95% CI = 1.25-1.42), PGS for Self-rated Health (Δ*R*^2^ = .014, ΔAUC = .021, OR = 0.75, 95% CI = 0.70-0.80), and PGS for neuroticism computed with LDAK (Δ*R*^2^ = .013, ΔAUC = .020, OR = 1.30, 95% CI = 1.22-1.39). The pattern of results was similar for the other outcomes, with DEP-PGS being the best or second-best predictor, but the predictive performance of PGSs was markedly reduced as indicated by lower *R*^2^ values and lower ORs per *SD* PGS ( Tables S3-S4 and Figure 2). The mean value of DEP-PGS (*z*-scores) in the Combined anxiety and depression group (mean = 0.33) was almost double the mean value in the Depression-only (mean = 0.17) and Anxiety-only (mean = 0.16) groups. For comparison, mean values were 0.09 in Medication-only, and –0.08 in the non-medicated control group. One-way ANOVA [*F*(4, 12787) = 54.63, *p* < 2e-16] followed by post-hoc tests, showed that DEP-PGS was on average significantly higher in each outcome group (Combined anxiety and depression, Anxiety-only, Depression-only, Medication-only) compared with the non-medicated control group. However, the Combined anxiety and depression group also had significantly higher mean DEP-PGS as compared to the other outcome groups, indicating that this is a disorder combination with higher polygenic load than anxiety or depression alone (see Supplemental Table S2 for results of post-hoc tests).

**Figure 2.**
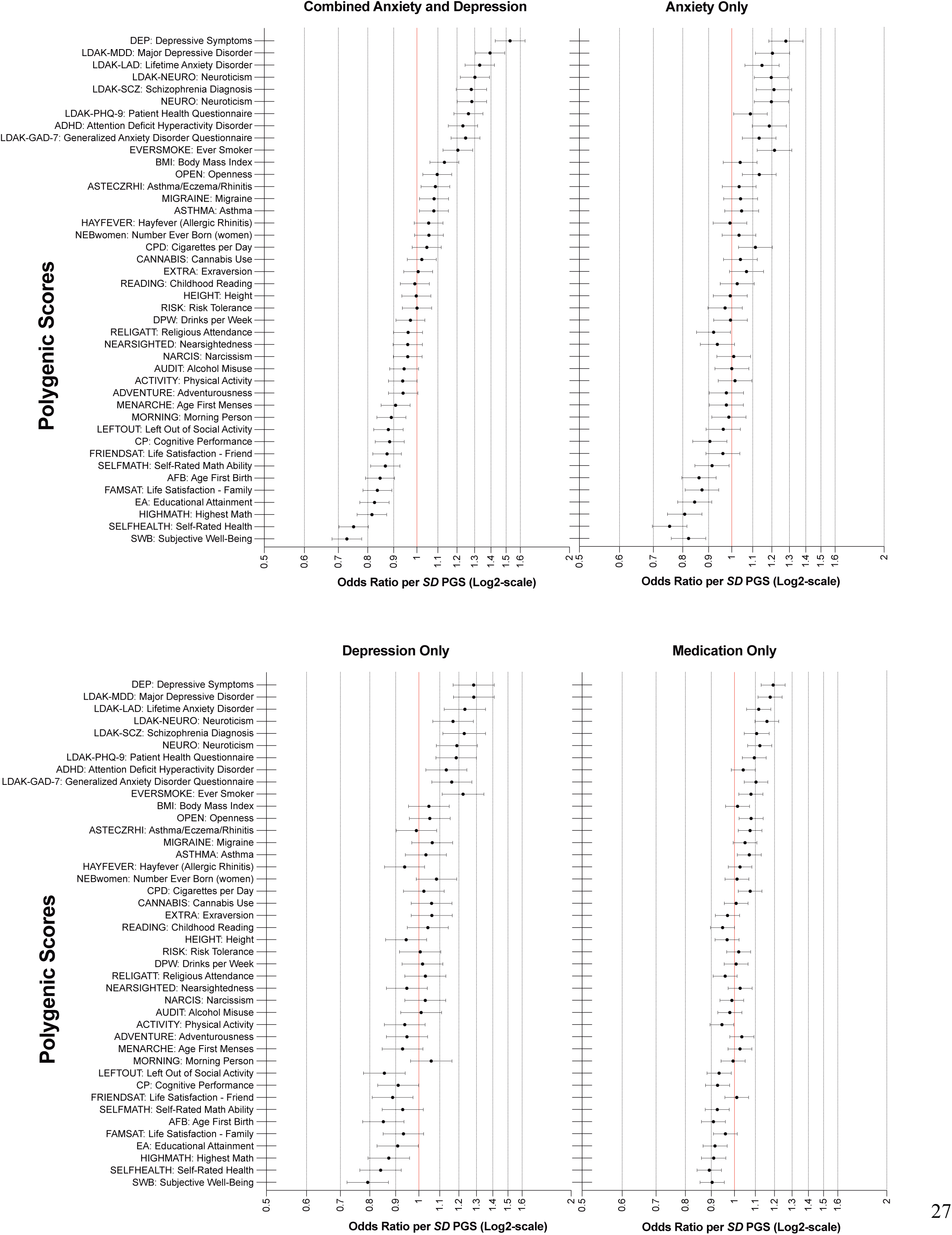
Adjusted OR per *SD* increase in PGSs predicting prescription-based outcomes. Error bars represent 95% CI.

### Clinical Diagnoses Regressed on Individual PGSs

To validate case classifications based on drug prescriptions, the regression analyses with PGSs as individual predictors were repeated, but this time with the clinical diagnosis groups as outcomes. For the combined depression and anxiety diagnoses group (base model *R*^2^ = .047, AUC = .667), DEP-PGS was the strongest predictor (Δ*R*^2^ = .036, ΔAUC = .051, OR = 1.70, 95% CI = 1.53-1.90) followed by SWB-PGS (Δ*R*^2^ = .030, ΔAUC = .048, OR = 0.61, 95% CI = 0.55-0.68) and MDD-PGS (Δ*R*^2^ = .026, ΔAUC = .038, OR = 1.59, 95% CI = 1.43-1.77). When predicting the probability of having an anxiety diagnosis (base model *R*^2^ = .032, AUC = .644), LAD-PGS (Δ*R*^2^ = .013, ΔAUC = .022, OR = 1.41, 95% CI = 1.25-1.60), DEP-PGS (Δ*R*^2^ = .013, ΔAUC = .016, OR = 1.40, 95% CI = 1.23-1.58) and MDD-PGS (Δ*R*^2^ = .012, ΔAUC = .018, OR = 1.39, 95% CI = 1.23-1.57) emerged as the top predictors. For the depression diagnosis group (base model *R*^2^ = .026, AUC = .631), DEP-PGS (Δ*R*^2^ = .015, ΔAUC = .028, OR = 1.41, 95% CI = 1.27-1.57), MDD-PGS (Δ*R*^2^ = .015, ΔAUC = .022, OR = 1.40, 95% CI = 1.26-1.55) and SWB-PGS (Δ*R*^2^ = .014, ΔAUC = .029, OR = 0.71, 95% CI = 0.64-0.79) were the PGSs with the strongest predictive performance (see suppl. Tables S5-S6 for OR, *R*^2^, *N*, and *p*-values for PGSs predicting ICD diagnoses outcomes). DEP-PGS had the overall highest accuracy and predicted more variation in the combined anxiety and depression diagnoses group compared with those having only one of these diagnoses, which is consistent with results from the regression analyses with symptom categories defined by drug prescription (see Figure 3). Hence, subsequent analyses include only prescription-based symptom categories as outcomes, since they include more cases than the diagnostically defined groups.

**Figure 3.**
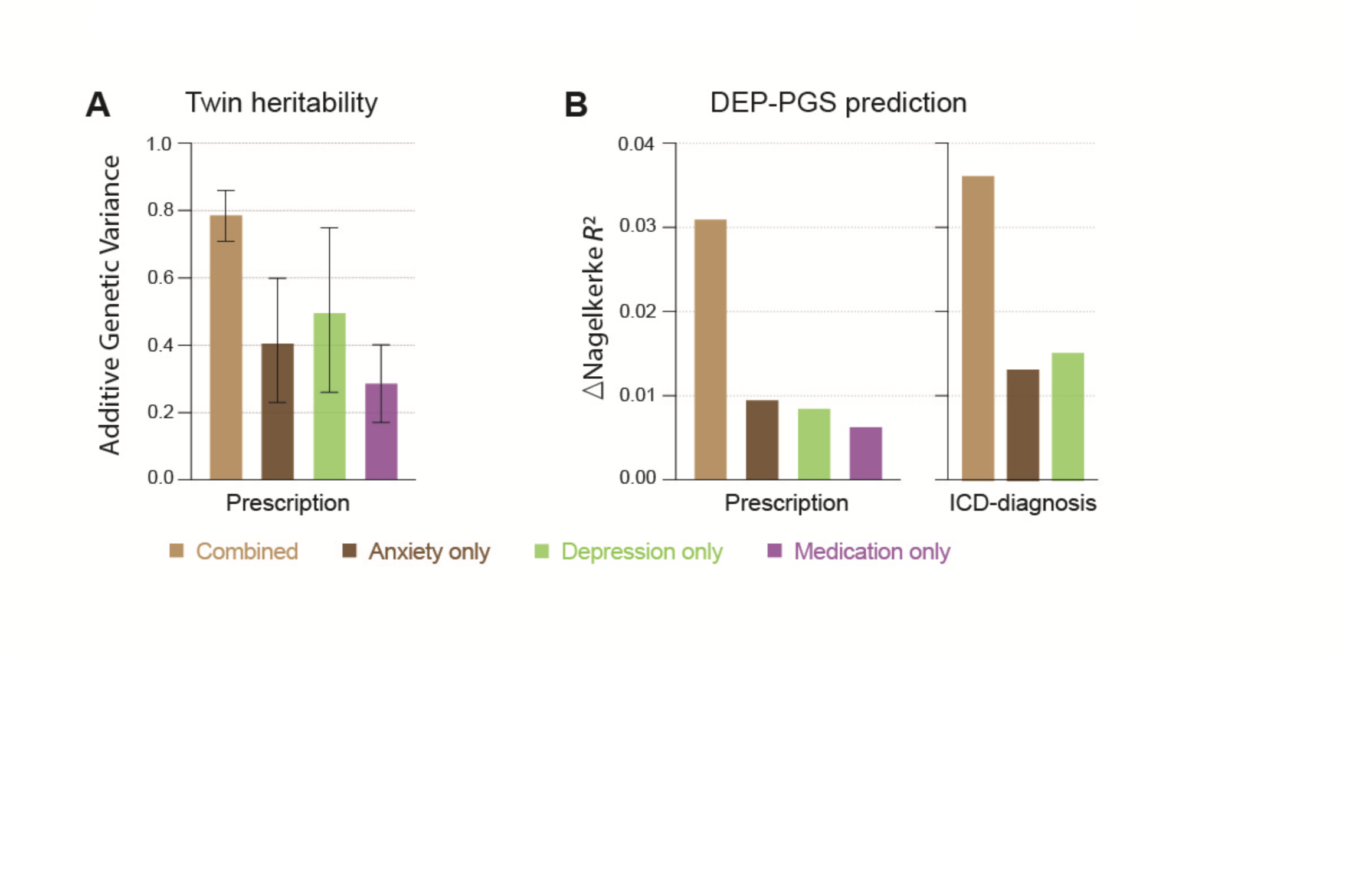
Genetic liability for anxiety and depression outcomes. (A) Twin heritability for each outcome (AE-model), error bars represent 95% CI. (B) Accuracy (Δ*R*^2^) of depressive symptoms PGS (DEP-PGS) from the Polygenic Index Repository in predicting each outcome above the baseline model.

### Regression models with multiple PGS predictors

In a single regression model containing all 42 PGSs simultaneously (base model *R*^2^ = .041, AUC = .632), only DEP-PGS (OR = 1.23, 95% CI = 1.12–1.35), SCZ-PGS (OR = 1.18, 95% CI = 1.10–1.27), and MDD-PGS (OR = 1.14, 95% CI = 1.06–1.24) remained significantly associated with drug prescription for Combined anxiety and depression (Δ*R*^2^ = .057, ΔAUC = .071). When predicting prescriptions for Anxiety-only (base model *R*^2^ = .023, AUC = .605), again using all 42 PGSs (Δ*R*^2^ = .034, ΔAUC = .057), the only significant predictors were SelfHealth-PGS (OR = 0.800, 95% CI = 0.72–0.89) and SCZ-PGS (OR = 1.15, 95% CI = 1.05–1.25). When predicting drug prescription for the Depression-only group (Δ*R*^2^ = .031, ΔAUC = .046) and the Medication-only group (Δ*R*^2^ = .017, ΔAUC = .025), no PGS-predictor survived the correction for multiple comparisons (see suppl. Tables S7-S8 for *R*^2^, OR and *p*-values). These results indicate that using multiple PGSs in the same model improves prediction compared to using individual PGSs, but that only a few PGSs have a unique contribution. Also, SCZ-PGS seem to have a unique contribution to the genetic influence on the Combined anxiety and depression and Anxiety-only categories although not being among the top 5 predictors in the regression models using single PGSs.

To check for possible confounding of zygosity, the sample was split by kinship and all regression analyses were repeated on the subsamples, with no substantial difference in the pattern of results compared to findings based on whole-sample analyses (see suppl. Tables S9-S16 for regression model estimates on split-half samples with no kinship).

### Outcome associations across the PGS distribution

We next wanted to know how extremely high or low PGSs predict drug prescription, as this could indicate whether scores could be useful to identify groups of vulnerable individuals. The highest and lowest deciles of DEP-PGS, the overall best performing genetic index, were compared with the middle 80% of the sample to estimate ORs. Individuals in the top 10% had greater odds of being prescribed medication for Combined anxiety and depressive symptoms (OR = 2.02, CI95% = 1.67-2.42), while those in the bottom 10% had a decreased risk (OR = 0.37, CI95% = 0.27–0.51). Hence, individuals with a PGS in one of the extreme ends of the polygenic continuum carry genetic risk or resilience compared to the rest of the distribution (see regression model estimates for all outcomes in suppl. Table S17).

To evaluate whether polygenic risk increased linearly across deciles in the distribution, we computed the OR for each decile compared to the lowest decile. We found a cumulative increase in OR with each decile for DEP-PGS (Figure 4 and suppl. Table S18). These results confirm to expectations from an additive infinitesimal model. The largest OR was found when comparing the lowest and highest deciles of the DEP-PGS (OR = 5.35, 95% CI = 3.78-7.73) in predicting prescription-based symptoms of Combined anxiety and depression.

**Figure 4.**
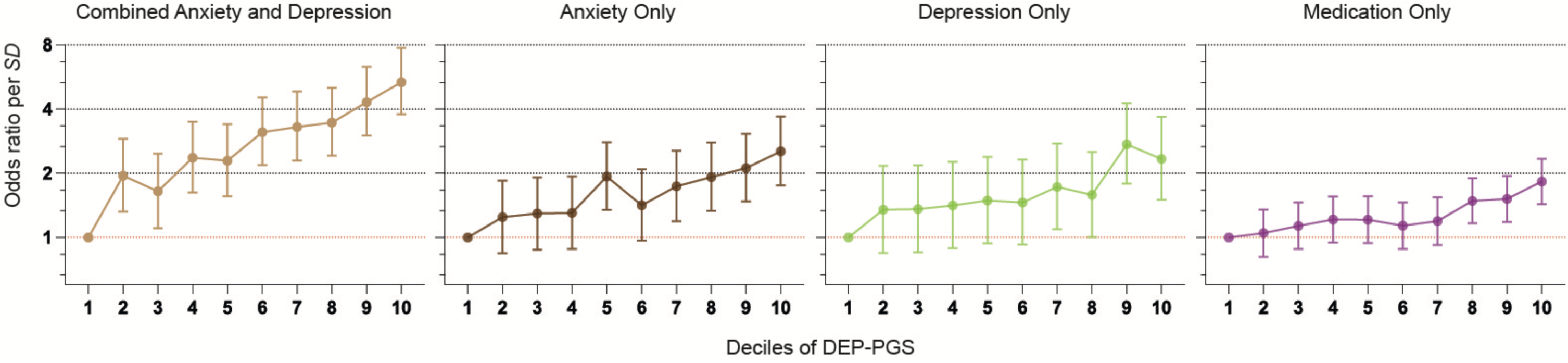
Genetic risk across the PGS distribution. OR per *SD* PGS with 95% CI for each decile of Depressive symptoms PGS (DEP-PGS) compared with the first decile.

## Discussion

Using polygenic scores and classical twin models, we investigated genetic influences on prescription of antidepressant and anxiolytic drugs for combined anxiety and depression relative to indications of anxiety or depressive symptoms only. Heritability was 79% for combined anxiety and depression, which was larger than for anxiety or depression alone, and is on par with heritability estimates for schizophrenia (39) and bipolar disorder (40). Combined anxiety and depression was also the outcome most strongly associated with PGSs, indicating that SNP-based prediction concurred with twin-based heritability estimates. These results were corroborated by PGSs more accurately predicting combined clinical depression and anxiety diagnoses compared with those having only a depression or an anxiety diagnosis. Thus, the results from two complementary methods to evaluate genetic influence, and two different case classification approaches, indicate a stronger genetic influence on the Combined symptom category than on anxiety or depressive symptoms alone.

It is well-known that the combination of anxiety and depressive symptoms is more impairing and difficult to treat than each symptom category alone (7–9). Despite this knowledge, the aetiology of this phenotype combination is poorly understood, and our study represents one of the few attempts made to investigate its genetic basis. There is evidence that the shared genetic component between anxiety and depression is substantially larger than the unique contributions to any of these phenotypes on their own. Large scale GWASs have found robust genetic correlations in the range of 80-95% (16,17), with 99% of risk variants for anxiety also influencing depression and 73% of risk variants for depression also conferring liability to anxiety (21). A conclusion from these findings is that the genetic influence on anxiety and depression is best explained by a common genetic factor. Accordingly, the fraction of unique variants that influence each phenotype could, when combined, only represent a small portion of the increase in genetic liability between the Combined anxiety and depression outcome compared to Anxiety– or Depression-only outcomes. An important consideration is that the large genetic overlap between anxiety and depression has been estimated in GWASs employing minimal phenotyping approaches that do not consider phenotypic heterogeneity (18). Recent findings based on more refined phenotyping showed that MDD with comorbid anxiety was twice as heritable compared to a broadly defined depression phenotype (41), indicating that there may be unique subsets of genetic variants associated with certain disorder subtypes not captured by previous GWASs. Thus, performing a large scale GWASs of the Combined anxiety and depression symptom category could be of value, as it would probably increase the predictive power of PGS predictions and help to tease out whether loci associated with this phenotype combination differs from MDD or anxiety disorders alone.

In line with the reported large genetic correlations between anxiety and depression (16,17,21), PGS for depressive symptoms predicted Anxiety-only to the same extent as Depression-only. The same pattern emerged for the PGS for lifetime anxiety diagnosis (LAD-PGS) that did not predict anxiety better than depression. PGSs for psychiatric disorders (anxiety, depression, schizophrenia, and ADHD) were however better at predicting anxiety, depression and their combination than were PGSs for physical disorders, body length or consumption of alcohol or marijuana. PGSs for subjective wellbeing and cognitive measures in turn predicted resilience. When controlling for all PGSs, the PGS for schizophrenia emerged as a significant predictor for the Anxiety-only and Combined symptom categories, suggesting that this PGS includes unique risk variants common to several disorders.

The finding of a distinction between the genetic contribution to combined anxiety and depressive symptoms relative to their isolated manifestations could reflect a more general genetic liability for psychopathology. In a meta-analysis across eight disorders, the Cross Disorder Group of the Psychiatric Genomics Consortium found 109 pleiotropic loci associated with two or more disorders (42), which extends previous results of shared common variant risk across schizophrenia, MDD, bipolar disorder, anxiety disorders, and ADHD (43). Thus, the high degree of genetic overlap among psychiatric disorders suggests that clinical classifications of discrete disorder categories do not reflect distinct underlying processes at the genetic level. A dimensional account of psychopathology has suggested that most common psychiatric disorders are unified by a single transdiagnostic dimension representing lesser-to-greater severity of psychopathology, referred to as the p-factor (44). Combined anxiety and depression represent greater severity than anxiety or depression alone, and the stronger genetic influence on the Combined anxiety and depression outcome could be a result of a genetic factor that makes individuals prone to developing psychiatric illness in general (45). Extending this line of thought to other psychiatric diagnoses, it could be predicted that the combination of any two diagnostic or symptom categories would be associated with greater heritability than each diagnosis in isolation. This hypothesis could be tested in future research.

Increasing the predictive performance of PGSs is important for risk stratification. We employed AUC estimates to index how well PGSs discriminated between participants with or without depression and anxiety. Participants with combined depression and anxiety determined from prescription data were distinguished from participants without depression or anxiety with an AUC of 68% (i.e., 18% above chance level of 50%) based on DEP-PGS, sex, age and ancestral principal components. The DEP-PGS by itself contributed 4.4% to the overall classification accuracy of the model. When Combined depression and anxiety was defined by ICD diagnoses instead of from prescription data, the AUC was 72% (5.1% unique contribution by DEP-PGS). An AUC of 70% is generally considered fair (46) and a 4-5% increase in overall accuracy solely based on one polygenic score is an important improvement in predictive performance, but questionable for the purpose of risk screening in the general population. To determine a cut-off in terms of AUC for when predictive factors should be implemented in screening and preventive treatment efforts is complex and depends on factors such as patient benefits, costs for the health care system and eventual risks associated with the intervention. For comparison, population screening for adverse childhood events, a well-known social risk factor for psychiatric disorders (47–49) is already underway in the USA where individuals with high scores on an adverse childhood events inventory are referred for health interventions (50). Adverse childhood event scores have been shown to predict anxiety with an AUC ranging from 54% to 59%, and depression with an AUC between 57% and 62% depending on whether adverse childhood events were assessed prospectively or retrospectively (51) which underscores that even modest AUCs can justify interventions if the patient benefits are judged to be large enough. Thus, if PGSs predicting anxiety and depression in the future also can be shown to predict responses to preventive interventions, they could be useful in guiding treatment decisions to improve patient health. We further foresee that PGSs are likely to complement other features in panels of prognostic markers as their predictive performance will increase.

The study had limitations. Depression related PGSs were based on larger discovery samples than anxiety-related scores, which impacts the results, as the out-of-sample predictive performance of a PGS is dependent on the definition of the phenotype and size of the discovery sample (20). Therefore, predictive performance will likely increase with larger samples and combined with more refined phenotyping. Outcomes were based on symptom categories and treated as life-time events. Consequently, we do not know if individuals’ anxiety and depression co-occurred or whether they were separated in time in the group with combined anxiety and depressive symptoms. In either case, our results point to the increased genetic propensity of having been prescribed drugs for combined anxiety and depressive symptoms or having been diagnosed with both disorders in a clinical setting.

In conclusion, we report larger genetic influence on combined anxiety and depressive symptoms than on either symptom category alone. A tentative explanation could be that depression and anxiety can be described by one latent variable and that increasing severity along this latent variable is associated with increased genetic liability.

## Data Availability

All data has been provided by the Swedish Twin Registry and any access needs to be approved and granted by them.

## Acknowledgments and Disclosures

This research was supported by grants from the Swedish Research Council (2018–01322) and the Bank of Sweden Tercentenary Foundation (P20-0125). The computations and data handling were enabled by resources provided by the National Academic Infrastructure for Supercomputing in Sweden (NAISS) and the Swedish National Infrastructure for Computing (SNIC) at Uppsala Multidisciplinary Center for Advanced Computational Science (UPPMAX) partially funded by the Swedish Research Council through grant agreements no. 2022-06725 and no. 2018-05973.

Preliminary analyses of the data reported here have been presented as abstract/poster at conferences.

Henrik Larsson reports receiving grants from Shire Pharmaceuticals; personal fees from and serving as a speaker for Medice, Shire/Takeda Pharmaceuticals and Evolan Pharma AB; and sponsorship for a conference on attention-deficit/hyperactivity disorder from Shire/Takeda Pharmaceuticals and Evolan Pharma AB, all outside the submitted work. Henrik Larsson is editor-in-chief of JCPP Advances. No other authors report financial interests or potential conflicts of interest.

## Author contributions

F.Å. designed the study. F.T. and J.R. performed the behavioral genetics analyses and ran the regression models, F.Å. contributed to interpretation of results. D.S. and X.Z. computed PGSs based on GWAS summary statistics. F.T. and F.Å. wrote the first draft. All authors substantially revised the manuscript. All authors have read and approved the manuscript.

## References

1. Bandelow B, Michaelis S. Epidemiology of anxiety disorders in the 21st century. Dialogues Clin Neurosci. 2015;17(3):327–35.

2. Lim GY, Tam WW, Lu Y, Ho CS, Zhang MW, Ho RC. Prevalence of Depression in the Community from 30 Countries between 1994 and 2014 /692/699/476/1414 /692/499 article. Sci Rep [Internet]. 2018;8(1):1–10. Available from: 10.1038/s41598-018-21243-x

3. Kessler RC, Gruber M, Hettema JM, Hwang I, Sampson N, Yonkers KA. Co-morbid major depression and generalized anxiety disorders in the National Comorbidity Survey follow-up. Psychol Med. 2008 Mar;38(3):365–74.

4. Hirschfeld RMA. The comorbidity of major depression and anxiety disorders: Recognition and management in primary care. Prim Care Companion J Clin Psychiatry. 2001;3(6):244–54.

5. Mineka S, Watson D, Clark LA. Comorbidity of anxiety and unipolar mood disorders. Annu Rev Psychol. 1998;49(February 1998):377–412.

6. Olfson M, Fireman B, Weissman MM, Leon AC, Sheehan D, Kathol RG, et al. Mental disorders and disability among patients in a primary care group practice. American Journal of Psychiatry [Internet]. 1997;154(12):1734–40. Available from: http://ovidsp.ovid.com/ovidweb.cgi?T=JS&PAGE=reference&D=emed7&NEWS=N&AN=27516592

7. Dold M, Bartova L, Souery D, Mendlewicz J, Serretti A, Porcelli S, et al. Clinical characteristics and treatment outcomes of patients with major depressive disorder and comorbid anxiety disorders – results from a European multicenter study. J Psychiatr Res [Internet]. 2017;91:1–13. Available from: 10.1016/j.jpsychires.2017.02.020

8. Fava M, Rush AJ, Alpert JE, Ph D, Balasubramani GK, Ph D, et al. Difference in Treatment Outcome in Outpatients With Anxious Versus Nonanxious Depression: A STAR * D Report. 2008;(March):342–51.

9. Tiller JWG. Depression and anxiety. Medical Journal of Australia. 2012;1(October):28–32.

10. Kendall KM, Van Assche E, Andlauer TFM, Choi KW, Luykx JJ, Schulte EC, et al. The genetic basis of major depression. Psychol Med [Internet]. 2021 Oct 8;51(13):2217–30. Available from: https://www.cambridge.org/core/product/identifier/S0033291721000441/type/journ al_article

11. Smoller JW. The Genetics of Stress-Related Disorders: PTSD, Depression, and Anxiety Disorders. Neuropsychopharmacology. 2016;41(1):297–319.

12. Middeldorp CM, Cath DC, Van Dyck R, Boomsma DI. The co-morbidity of anxiety and depression in the perspective of genetic epidemiology. A review of twin and family studies. Psychol Med. 2005;35(5):611–24.

13. Hettema JM. What is the genetic relationship between anxiety and depression? Vol. 148, American Journal of Medical Genetics, Part C: Seminars in Medical Genetics. 2008. p. 140–6.

14. Guffanti G, Gameroff MJ, Warner V, Talati A, Glatt CE, Wickramaratne P, et al. Heritability of major depressive and comorbid anxiety disorders in multi-generational families at high risk for depression. American Journal of Medical Genetics, Part B: Neuropsychiatric Genetics. 2016 Dec 1;171(8):1072–9.

15. Torkamani A, Wineinger NE, Topol EJ. The personal and clinical utility of polygenic risk scores. Nat Rev Genet [Internet]. 2018;19(9):581–90. Available from: 10.1038/s41576-018-0018-x

16. Levey DF, Gelernter J, Polimanti R, Zhou H, Cheng Z, Aslan M, et al. Reproducible Genetic Risk Loci for Anxiety: Results From ∼200,000 Participants in the Million Veteran Program. Am J Psychiatry. 2020;177(3):223–32.

17. Purves KL, Coleman JRI, Meier SM, Rayner C, Davis KAS, Cheesman R, et al. A major role for common genetic variation in anxiety disorders. Mol Psychiatry [Internet]. 2020;25(12):3292–303. Available from: 10.1038/s41380-019-0559-1

18. Cai N, Revez JA, Adams MJ, Andlauer TFM, Breen G, Byrne EM, et al. Minimal phenotyping yields genome-wide association signals of low specificity for major depression. Nat Genet [Internet]. Available from: 10.1038/s41588-020-0594-5

19. Wray NR, Ripke S, Mattheisen M, Trzaskowski M, Byrne EM, Abdellaoui A, et al. Genome-wide association analyses identify 44 risk variants and refine the genetic architecture of major depression. Nat Genet. 2018;50(5):668–81.

20. Mitchell BL, Thorp JG, Wu Y, Campos AI, Nyholt DR, Gordon SD, et al. Polygenic Risk Scores Derived From Varying Definitions of Depression and Risk of Depression. JAMA Psychiatry. 2021 Oct;78(10):1152–60.

21. Als TD, Kurki MI, Grove J, Voloudakis G, Therrien K, Tasanko E, et al. Depression pathophysiology, risk prediction of recurrence and comorbid psychiatric disorders using genome-wide analyses. Nat Med. 2023 Jul 1;29(7):1832–44.

22. Valderas JM, Starfield B, Sibbald B, Salisbury C, Roland M. Defining comorbidity: Implications for understanding health and health services. Ann Fam Med. 2009;7(4):357– 63.

23. Becker J, Burik CAP, Goldman G, Wang N, Jayashankar H, Bennett M, et al. Resource profile and user guide of the Polygenic Index Repository. Nat Hum Behav. 2021;

24. Lichtenstein P, Sullivan PF, Cnattingius S, Gatz M, Johansson S, Carlström E, et al. The Swedish Twin Registry in the third millennium: An update. Twin Research and Human Genetics. 2006;9(6):875–82.

25. Zagai U, Lichtenstein P, Pedersen NL, Magnusson PKE. The Swedish Twin Registry: Content and Management as a Research Infrastructure. Twin Research and Human Genetics. 2019;22(6):672–80.

26. Donahue KL, D’Onofrio BM, Lichtenstein P, Långström N. Testing Putative Causal Associations of Risk Factors for Early Intercourse in the Study of Twin Adults: Genes and Environment (STAGE). Arch Sex Behav. 2013 Jan 23;42(1):35–44.

27. Socialstyrelsen. National Prescribed Drug Register. . 2020 [cited 2023 Feb 27]. Socialstyrelsen. Available from: https://www.socialstyrelsen.se/en/statistics-and-data/registers/national-prescribed-drug-register/

28. WHO. WHO. Collaborating Centre for Drug Statistics Methodology [Internet]. 2021 [cited 2023 Feb 27]; Available from: https://www.whocc.no/

29. UK Biobank. Vol. 7, Protocol No: UKBB-PROT-09-06.PROT-09-06. 2007. p. 1–112 UK Biobank: Protocol for a large-scale prospective epidemiological resource. Available from: http://www.ukbiobank.ac.uk/wp-content/uploads/2011/11/UK-Biobank-Protocol.pdf.

30. Howard DM, Adams MJ, Clarke TK, Hafferty JD, Gibson J, Shirali M, et al. Genome-wide meta-analysis of depression identifies 102 independent variants and highlights the importance of the prefrontal brain regions. Nat Neurosci [Internet]. 2019;22(3):343–52. Available from: 10.1038/s41593-018-0326-7

31. Ripke S, Walters JTR, O’Donovan MC, Consortium TSWG of the PG. Mapping genomic loci prioritises genes and implicates synaptic biology in schizophrenia. medRxiv [Internet]. 2020 Jan 1;2020.09.12.20192922. Available from: http://medrxiv.org/content/early/2020/09/13/2020.09.12.20192922.abstract

32. Sudlow C, Gallacher J, Allen N, Beral V, Burton P, Danesh J, et al. UK Biobank: An Open Access Resource for Identifying the Causes of a Wide Range of Complex Diseases of Middle and Old Age. PLoS Med. 2015;12(3):1–10.

33. Speed D, Holmes J, Balding DJ. Evaluating and improving heritability models using summary statistics. Nat Genet. 2020 Apr 23;52(4):458–62.

34. Zhang Q, Privé F, Vilhjálmsson B, Speed D. Improved genetic prediction of complex traits from individual-level data or summary statistics. Nat Commun [Internet]. 2021;12(1):1–9. Available from: 10.1038/s41467-021-24485-y

35. Falconer DS. The inheritance of liability to certain diseases, estimated from the incidence among relatives. Ann Hum Genet. 1965;29.

36. Neale MC, Maes HH. Methodology for Genetic Studies of Twins and Families. Dordrecht: Kluwer Academics. 2004;

37. Holst KK, Scheike T. mets (R package). Version 0.2.8. . URL: http://cran.r-project.org/web/packages/mets/.

38. Benjamini Y, Hochberg Y. Controlling the False Discovery Rate: A Practical and Powerful Approach to Multiple Testing. Journal of the Royal Statistical Society: Series B (Methodological). 1995;57(1):289–300.

39. Hilker R, Helenius D, Fagerlund B, Skytthe A, Christensen K, Werge TM, et al. Heritability of Schizophrenia and Schizophrenia Spectrum Based on the Nationwide Danish Twin Register. Biol Psychiatry. 2018 Mar;83(6):492–8.

40. Gordovez FJA, McMahon FJ. The genetics of bipolar disorder. Mol Psychiatry. 2020 Mar;25(3):544–59.

41. Nguyen TD, Harder A, Xiong Y, Kowalec K, Hägg S, Cai N, et al. Genetic heterogeneity and subtypes of major depression. Mol Psychiatry [Internet]. 2022 Mar 8;27(3):1667–75. Available from: https://www.nature.com/articles/s41380-021-01413-6

42. Lee PH, Anttila V, Won H, Feng YCA, Rosenthal J, Zhu Z, et al. Genomic Relationships, Novel Loci, and Pleiotropic Mechanisms across Eight Psychiatric Disorders. Cell [Internet]. 2019 Dec;179(7):1469–1482.e11. Available from: https://linkinghub.elsevier.com/retrieve/pii/S0092867419312760

43. Anttila V, Bulik-Sullivan B, Finucane HK, Walters RK, Bras J, Duncan L, et al. Analysis of shared heritability in common disorders of the brain. Science (1979). 2018 Jun 22;360(6395).

44. Caspi A, Houts RM, Belsky DW, Goldman-Mellor SJ, Harrington H, Israel S, et al. The p factor: One general psychopathology factor in the structure of psychiatric disorders? Clinical Psychological Science. 2014 Mar 1;2(2):119–37.

45. Waszczuk MA, Eaton NR, Krueger RF, Shackman AJ, Waldman ID, Zald DH, et al. Redefining Phenotypes to Advance Psychiatric Genetics: Implications From Hierarchical Taxonomy of Psychopathology. J Abnorm Psychol. 2019;

46. Rice ME, Harris GT. Comparing effect sizes in follow-up studies: ROC area, Cohen’s d, and r. Law Hum Behav. 2005 Oct;29(5):615–20.

47. Hughes K, Bellis MA, Hardcastle KA, Sethi D, Butchart A, Mikton C, et al. The effect of multiple adverse childhood experiences on health: a systematic review and meta-analysis. Lancet Public Health. 2017 Aug 1;2(8):e356–66.

48. Reuben A, Moffitt TE, Caspi A, Belsky DW, Harrington H, Schroeder F, et al. Lest we forget: comparing retrospective and prospective assessments of adverse childhood experiences in the prediction of adult health. J Child Psychol Psychiatry. 2016 Oct 1;57(10):1103–12.

49. Felitti VJ, Anda RF, Nordenberg D, Williamson DF, Spitz AM, Edwards V, et al. Relationship of childhood abuse and household dysfunction to many of the leading causes of death in adults: The adverse childhood experiences (ACE) study. Am J Prev Med. 1998 May;14(4):245–58.

50. Centers for Disease Control and Prevention. https://www.cdc.gov/violenceprevention/aces/ace-brfss.html. 2024. Behavioral Risk Factor Surveillance System Survey ACE Data, 2009-2021.

51. Baldwin JR, Caspi A, Meehan AJ, Ambler A, Arseneault L, Fisher HL, et al. Population vs Individual Prediction of Poor Health from Results of Adverse Childhood Experiences Screening. JAMA Pediatr. 2021 Apr 1;175(4):385–93.

